# Stricter protocols combined with a clinical serum biomarker can increase replicability and causality for dietary intervention studies. Plus empirical data on BPA regrettable substitutions

**DOI:** 10.1101/2022.08.09.22278588

**Authors:** W. Lewis Perdue, Victor I. Reus, Richard B. van Breemen, Ruth N. Muchiri, Rebecca L. Yeamans-Irwin

## Abstract

Effective regulation of harmful environmental chemicals found in wide variety of consumer products and consumables has been thwarted by the lack of agreement between government scientists and university/academic laboratories regarding the quantification of significant human harms. This is particularly relevant regarding plastic-derived chemicals (PDCs), such as Bisphenol A, now that the federal CLARITY-BPA program has failed to achieve any credible, human-significant scientific consensus. Because of this disagreement, direct, clinical human experimental data is vital to resolving this situation. In an effort to develop direct human-relevant data, some academic investigators have employed dietary intervention studies in an attempt to shed light on the controversy. Unfortunately, dietary intervention efforts thus far have not demonstrated causality or replicability.

Investigators of this study propose a novel human dietary intervention protocol that can be both replicable and causal. This first-of-a-kind dietary intervention study explores a potential causal relationship between human serum levels of BPA and High-Sensitivity C-Reactive Protein (hsCRP), a proven clinical indicator of inflammation. Investigators used the equivalent of a USDA-defined typical diet followed by a PDC-reduced diet to compare blood levels of hsCRP. This proof-of-concept investigation is the first to use an easily accessible, medically-accepted clinical laboratory test to directly measure human health effects of PDC reduction.

Unexpected phenomena discovered during the investigation offer study protocol modifications to enhance widespread replicability, and economically practical expansion to a substantial proportion of the approximately 84,000 mostly unregulated chemicals found in the human environment. In addition, our LC/MS-MS results offer the first direct quantitative human clinical evidence (of which we are aware) confirming the existence of regrettable substitutions in which product manufacturers have reduced BPA usage while substituting Bisphenol analogues that appear to equal or exceed BPA human toxicity. Bolstered by the unexpected results in this proof-of-concept investigation, novel lessons and techniques described herein may further specific and improved methods and best practices that can enable future dietary interventions to produce replicable, causal human results.

## BACKGROUND

Exposure to environmental chemicals in the U.S. is widespread.^1^ As of June 2020, more than 86,000 chemicals were approved for use in the United States,^2^ and at least 4,000 of those are Plastic-Derived Chemicals (PDCs) present in food contact materials.^3,4,5^ PDCs such as Bisphenol A (BPA), phthalates, and other plastic derivations are present in approximately 97% of the U.S. population.^6,7^ Human and animal studies have also identified PDCs compounds as contributors to cancer^10-21^, cardiovascular disorders^12,22-29,^ obesity^30-36^, type 2 diabetes^35,37-39^, metabolic syndrome^31-33,40,41^, neurological and behavioral disorders^42,43^ also including Alzheimer’s Disease^12,40,44-48^, as well as reproductive^13,49-56^, and developmental disorders^13,57-62^ and allergies^63-70^. PDCs such as BPA are classified as endocrine disrupters, even in low-level concentrations.^7,9^ More significantly, in a 10-year observational study of 3,883 adults in the United States, participants with higher urinary Bisphenol A levels were at a 49% greater risk for death from all causes.^8^

No clear scientific agreement exists about the nature and degree of health effects of most of these PDCs^71^. Even BPA, singled out for special attention by the U.S. government CLARITY-BPA program^72^ because of its ubiquity, failed to resolve a contentious scientific divide on safety. That divide persists as regulatory investigators have deemed current exposure levels as harmless^73-75^ while academic biomedical scientists continue to disagree and find BPA a public health threat.^76-79^

### Routes of Exposure

Exposure to BPA and other PDCs come from both dietary and non-dietary sources.^80^

BPA and other PDCs are found in household products such as detergents, cosmetics, lotions, and fragrances^81^, as well as in water bottles and baby bottles, thermal paper for printers, dental sealants, and medical devices including intravenous fluid and chemotherapy bags and tubing.^7,15,82-86^

Food and beverage packaging are substantial contributors to PDC physiological burdens.^7,87-91^ Consumers are exposed to many PDCs from leaching and migration of chemicals from plastics and other food contact materials.^7,88-96^

Exposure routes for food and beverages also include preservatives, flavorings, scents, texture enhancers, and coloring agents^5^, migration/leaching of chemicals from harvesting and processing^87^, as well as home food-handling which can accelerate migration through heating, microwaving, ultraviolet light exposure (including fluorescent lighting), and the contact of contaminated oils and alcohols with plastics.

Ultra-processed foods (UPFs) that have undergone substantial chemical or physical modification through manufacturing methods also have numerous avenues of PDC contamination due to residual contamination in additives, from additional contacts with plastic components, and from the addition of fats which facilitate the transfer of lipid-soluble polymer components into the UPFs.^98,99^

UPFs and PDCs are both associated with many non-communicable dietary-related diseases and syndromes including obesity,^100-107^ diabetes,^108-110^ cardiovascular disease,^111-113^ and cancer^10,18,114,115^.

### Could standard human clinical blood tests resolve the clarity issues with CLARITY?

CLARITY-BPA was established in 2014 as a joint effort by the Food and Drug Administration, National Institutes of Health, and the National Toxicology Program. The murine-based program, which has been conducted twice with rats accidentally contaminated with BPA prior to the trials, failed to reconcile differences, and has resulted in public acrimony between government and academic teams. Government teams using legacy regulatory methods insensitive to BPA have vocally accused academic teams of using state-of-the art techniques – such as those currently employed to develop new pharmaceuticals – that are not currently codified in government regulations. Academic teams assert that the legacy toxicology protocols – known as GLP (Good Laboratory Practices) are not sensitive enough to detect endocrinal, hormonal, and genetic impacts.

As a result, the current CLARITY-BPA debate centers on experimental protocol flaws, contrasting interpretations of published science, confounding factors, sources of contamination and other fine points of scientific experimental design as well as practice and fundamental differences concerning the existence of non-monotonic behavior of substances present at very low levels.

The CLARITY program has been criticized as irrelevant due to the failures of translating murine results to humans.^116^ Indeed, other published reports find that murine model results do not to translate accurately to humans as much as 92% of the time.^117^ Even pre-clinical research trials fail to be replicable^118^ from 51% to 89% of the time.^119-120^

Human dietary intervention studies, which might be able to shed light on the issue, have replication and scientific rigor flaws that prevent them from offering reliable clinical conclusions.^121,122^

Interpreting these factors by lay audiences can be all but impossible. Likewise, the current internecine arguments among scientists are unlikely to sway public, governmental or manufacturing opinion.

### Enter the humble clinical blood test

Significantly, a source of reproducible, causally connected, and clinically valuable direct human data may lurk in standard laboratory blood profiles.

Because of the role of inflammation in many BPA-associated non-communicable diseases, High-Sensitivity C-Reactive Protein (hsCRP) may fill the need for clinical insights into numerous PDC-linked inflammation-linked conditions cardiovascular disease,^123-125,131,137^ Type 2 Diabetes,^126-127^ cancer,^126,129,137^Alzheimer’s Disease,^130, 132^ depression, and suicide,^133-136^ and auto-immune diseases^137^ including IBD,^138^ rheumatoid arthritis,^139^ and lupus.^140^

In addition, BPA has been found to activate the same NLRP3 inflammasome pathway activity^141^ implicated in cytokine storms which develop in research suspects along with bradykinin disorders^142^ as causes underpinning some of the most serious of COVID-19 cases.^143-145^

### OBJECTIVES

**Objective 1, Marker Validation:** Can hsCRP serve as a marker for BPA exposures?

**Objective 2, Duration Evaluation:** Is a short (e.g. six-day) trial with three days each for contamination and intervention sufficient to affect measurable hsCRP outcomes?

**Objective 3, Verify Causality and Improve Replication:** Can replicability and causality of dietary intervention trials be increased by developing best practices and applying the discipline of standard laboratory practices to the sourcing, preparation, and serving of human food along with their complete data capture and reporting?

## METHODS

Given that the use of clinical blood tests as possible direct human health effects indicators of environmental chemical contamination is an unknown field, investigators felt that a small proof-of-concept trial (n-of-1) would be sufficient to test the validity of the concept and protocols before beginning a larger, more expensive study. N-of-1 investigations have found wide acceptance and success in biomedical science^146^ and are proposed as a main tactic in precision medicine^147^ and pharmaceutical research.

This n-of-1, six-day study (SSHS) was approved by the Committee on Human Research/IRB at the University of California San Francisco School of Medicine.^148^ The study consisted of two legs - a three-day typical American diet (Typical) with known sources of plastic contamination followed by a three-day intervention diet (Intervention) of foods with measurably reduced BPA contamination.

This investigation was conducted in a 400-square-foot professional kitchen supervised and directed by investigators in the same manner as a bench lab experiment. Central air with HEPA filters ventilated the kitchen which was also equipped with two standalone HEPA filter machines. A 1,200 cubic-feet/minute, four-centrifugal-fan exhaust hoods were used over an eight-burner natural gas stove. Two electric ovens were used for baking.

Plastic items including utensils, containers, and cookware with non-stick coatings were removed from the kitchen prior to trial onset. Food preparation took place on stainless steel countertops. Kitchen-ready scales were obtained and calibrated using brass gram weights.

Indoor kitchen air quality (via PM2.5 levels) were monitored via sensors from PurpleAir.Com and maintained at an Air Quality Index of 0. Test subject consumed meals in an adjoining space with identical environmental conditions including the absence of plastic.

### A. Menu Determination

The menu for each of the two diets (Typical and Intervention) was made as identical as feasible.

#### Typical

The typical American diet (i.e. PDC-contaminated diet) was drawn from the United States Department of Agriculture (USDA).^149^ Food for the typical diet was sourced from national brands available at a large national chain store (Safeway) to maximize availability.

#### Intervention

For the intervention diet, the menu from typical American diet was adapted to offer replicate food items with reduced or absent PDC contamination.

Intervention food was sourced in accordance with Appendix 2 of the revised protocol.^148^

The extensive requirements of the revised protocol included sourcing food close as possible to its actual production from a vendor capable of shipping nationally. Additionally, the producer:

- Either dry-farmed or used well water for irrigation,
- Did not irrigate with recycled wastewater or biosolids (sewage plant sludge)
- Adhered -- as a minimum -- to USDA organic standards.

Because milk is sourced locally or regionally, even by large chains, milk samples were analyzed via LC-MS/MS performed by Eurofins (https://www.eurofins.com/) to ensure the same or similar BPA quantification. Cheese products were selected from nationally available dairy brands whose milk scored below the limit of quantification (LOQ) in the LC-MS/MS tests.

Fish and seafood were excluded because the aquatic environmental pollution variability made it impossible for consistency in BPA exposure.

Carbon-filtered water was used for food preparation and to rinse before use of all dishes, utensils, pots, pans, and food-contact appliances.

Multi-ingredient foods, such as a spaghetti and meatball frozen entrees, were reverse engineered from the dishes used in the contamination leg. Special attention was devoted to the health effects of micro- and nanoplastics, and to minimizing exposure to those particles of undeterminable composition.

### B. Food Preparation

After lengthy research, this study’s protocols were extensively revised^148^ to include the development of best practices that included the imposition of specific, rigorous scientific practices common to bench laboratory investigations. This included rigorous requirements for sourcing food and beverages, and the realization that a kitchen needed to be treated as a proper laboratory environment.

These constraints required investigators to train and supervise all kitchen personnel to assure proficiency in precise measurements, and treating ingredients as reagents, recipes as procedures, appliances as equipment, and the paramount requirement to capture detailed and extensive data capture sufficient to allow replication of the trial under the exact same conditions. The most proficient were those with substantial baking experience.

In the kitchen, only glass, stainless steel, aluminum foil, and tight grain maple cutting boards were permitted for food preparation. 100% cotton dish towels were allowed; paper towels could only be used for cleanup. Nitrile gloves were used for all food preparation.

Intensive treatment of food in the revised protocol^148^ was designed to minimize BPA. For example, fresh organic vegetables were thoroughly rinsed or soaked in filtered water. Solid meat was first wiped with extra virgin olive oil obtained from a local mill which uses no plastic and packs the oil in glass. The meat was then scraped with a metal pastry divider and wiped dry again to remove lipid-solid PDCs such as BPA. Hamburger was prepared fresh using a grinder with all metal food contact surfaces.

No pre-prepared foods were allowed. Pasta was made and bread was baked using flour from a small mill that grinds wheat with no plastic contact obtained from small local organic, pesticide-free farms.

All measurements (including for liquids) were made by metric weight. All main ingredients were measured to the nearest gram, but small-weight items such as spices, herbs, other seasonings, and other items were measured to the nearest tenth of a gram.

Seasoning was with whole spices and fresh herbs with no PDC exposure grown for the purpose of this study by the investigators. When investigator-grown seasonings were not available, investigators specified organic whole spices (cinnamon sticks, whole nutmeg fruit, peppercorns) and hand ground them after wiping or rinsing with filtered water. Items with known or suspected plastic contamination (such as micro-plastic contamination of table salt^150^ were replaced with suitable reagents from Sigma-Aldrich.

### C. Blood Sampling of Study Participants

Two blood samples (one for hsCRP and one for BPA levels) were taken from the subject three times during the study: the first day to establish baseline; the last morning after the typical menu leg; and the last morning after the intervention leg.

All blood draws were made at Sonoma Valley Hospital (SVH - an affiliate of UCSF).

Samples to be analyzed for hsCRP were drawn in light-green-topped tubes with standard concentrations of Li-heparin and centrifuged. Those were analyzed by the UCSF Parnassus campus medical laboratory.

The samples for BPA analysis were drawn by SVH using a vacutainer kit composed of polymers tested by Dr. Roy Gerona’s UCSF which had been analyzed and found not to leach BPA or other PDC.

The virgin whole blood samples were hand-delivered within four hours of blood draws to Dr. Gerona’s UCSF research lab where they were centrifuged and frozen to -80C.

### LC-MS/MS Mass Spectrometry Analysis

The -80 cold chain was maintained for the samples which were later delivered to the Linus Pauling Institute at Oregon State University in Corvallis, OR There, the samples were analyzed using ultrahigh pressure liquid chromatography-tandem mass spectrometry using a triple quadrupole Shimadzu mass spectrometer which allows the study of the presence of other chemical compounds.

The lab also used a high-resolution quadrupole time-of-flight mass spectrometer using untargeted scanning for BPA, BPF and BPS which may be studied for the presence of other compounds.

See Supplementary Information for details.

## RESULTS

Levels of hsCRP decreased 21.4% from baseline to end of the Intervention leg: 1.1 mg/L from 1.4mg/L. The results demonstrated a final percentage reduction in hsCRP roughly parallel to (but smaller in magnitude) than that of a major NIH-funded dietary intervention by Hall, et al.^105^ published in 2019 approximately four months prior to our trial. (See Table 1)

**Table 1.** SSHS Study Results Versus Hall et al

| NIH/Hall versus SSHS |  |  |  |  |
| --- | --- | --- | --- | --- |
| hsCRP (mg/L) |  |  |  |  |
|  | NIH/Hall | leg delta | SSHS | leg delta |
| baseline | 2.7 |  | 1.4 |  |
|  |  | -0.3 |  | -0.5 |
| contamination | 2.4 |  | 0.9 |  |
|  |  | -0.9 |  | 0.2 |
| intervention | 1.5 |  | 1.1 |  |
| Total Delta |  | -1.2 |  | -0.3 |
| Total Delta % | -44.4 |  | -21.4 |  |

## DISCUSSION

This investigation’s primary goal was to determine whether inflammation levels as indicated by hsCRP would be sensitive enough to serve as a human clinical biomarker for Bisphenol A, and possibly other plastic derived chemicals. Because, at the time of this study, that relationship had not been previously established, an n-of-one cohort was chosen to avoid the expense of a larger cohort in the event that hsCRP was not sufficiently sensitive to the changes in levels of serum BPA resulting from dietary exposure.

This study was approved by UCSF-IRB on Nov. 15, 2014. That original IRB approval was substantially revised^148^ because the original protocol was patterned after existing published dietary interventions,^73, 89,105, 150,151^ whose designs were not capable of producing replicable results or credible causal relationships.^121,152^

While our data indicated an overall reduction in hsCRP from baseline to intervention, the data unexpectedly indicated a decrease at the end of the contamination leg.

The lower decline in hsCRP levels in our study versus Hall et al. could have been caused by our shorter trial duration (six days rather than 28) which affects the release of BPA from adipose stores and/or its metabolism. Indeed, the minor increase of hsCRP in the second leg of the trial may indeed be due to the delayed release of BPA from the test subjects’ dietary changes or relatively lower baseline BPA concentrations.

This could also be due to the fact that BPA may have a longer half-life than previously thought.^154^

An alternate explanation may lie in the known non-monotonic behavior of BPA, in which its effects become more powerful as the concentration falls into a specific range of influence.^155,156^

Non-monotonic behavior is counter-intuitive, but found in other compounds especially those that, like Bisphenol A, exhibit estrogenic effects active in the endocrine system. Breast-cancer treatment tamoxifen, for example, exhibits non-monotonic behavior and is most effective in tiny concentrations.^156^

Another significant difference between this trial and Hall et al., is that our trial was interrupted by Northern California wildfires that dramatically increased PM2.5 particulate pollution, a notorious promoter of inflammation.^158^ This exposed the n-of-1 investigator/test to environmental PM2.5 pollution in order to deliver blood draw samples to two UCSF laboratories at the Parnassus and Mt. Zion campuses. Those exposures lasted for a minimum of 2.5 hours on three occasions. That level of exposure may account for the overall smaller decrease in hsCRP in our study versus Hall et al.

### Bisphenol A levels below the Lower Level of Quantification

Mass spectrometry analysis indicated Bisphenol A levels below the Lower Level Of Quantification (LLOQ). See Table 2.

**Table 2.**

| Total Bisphenol + Observations at a Glance |  |  |  |  |  |  |
| --- | --- | --- | --- | --- | --- | --- |
| MG = monoglucuronide DG = diglucuronide. All concentrations, (ng/mL) |  |  |  |  |  |  |
| Study Segment | BPA (1) | BPA-MG (2) | BPF (3) | BPS (4) | BPF-MG (5) | BPS-DG (6) |
| BASELINE | <LLOQ TND | <LLOQ TND | 59.4 | 10.5 | 17.5 | 15.7 |
| Total Bisphenol |  |  | 69.9 |  | 33.2 |  |
| hsCRP(mg/L) |  |  | 1.4 |  |  |  |
| CONTAMINATION | <LLOQ TND | <LLOQ TND | 107 | 11.5 | 16 | 7.4 |
| Total Bisphenol |  |  | 118.5 |  | 23.4 |  |
| hsCRP(mg/L) |  |  | 0.9* |  |  |  |
| INTERVENTION | <LLOQ TND | <LLOQ TND | 63.9 | 11.1 | 28.8 | 4.8 |
| Total Bisphenol |  |  | 75 |  | 33.6 |  |
| hsCRP(mg/L) |  |  | 1.1 |  |  |  |
| <LLOQ TND: Below Lower Limit of Detection. Target Not Detected. |  |  |  |  |  |  |
| * hsCRP unexpectedly decreased during contamination segment. Overall intevention decrease from baseline. |  |  |  |  |  |  |
| 1. Below <LLOQ> (0.05) - Contrary to published studies which indicate ubiquitos BPA levels in U.S. population. |  |  |  |  |  |  |
| 2. NHANES data indicatates U.S. population has consistnt levels of BPA-G. |  |  |  |  |  |  |
| 3. BPF approximately the same for Baseline and Intervention. |  |  |  |  |  |  |
| 4. BPS approximately same for all segments. |  |  |  |  |  |  |
| 5. BPF-MG approximately same for baseline and contamination legs. Higher after intervention. |  |  |  |  |  |  |
| 6. BPS-DG higher in baseline than contamination. Substantially lower in intevention. |  |  |  |  |  |  |

This was not expected. Further, those results run contrary to NHANES published studies (See references Table 3) which indicate ubiquitous BPA-monoglucuronide levels in U.S. population. Because BPA-MG is the primary metabolic conjugate of BPA, the precursory compound – BPA – is assumed by the presence of BPA-MG

**Table 3.**
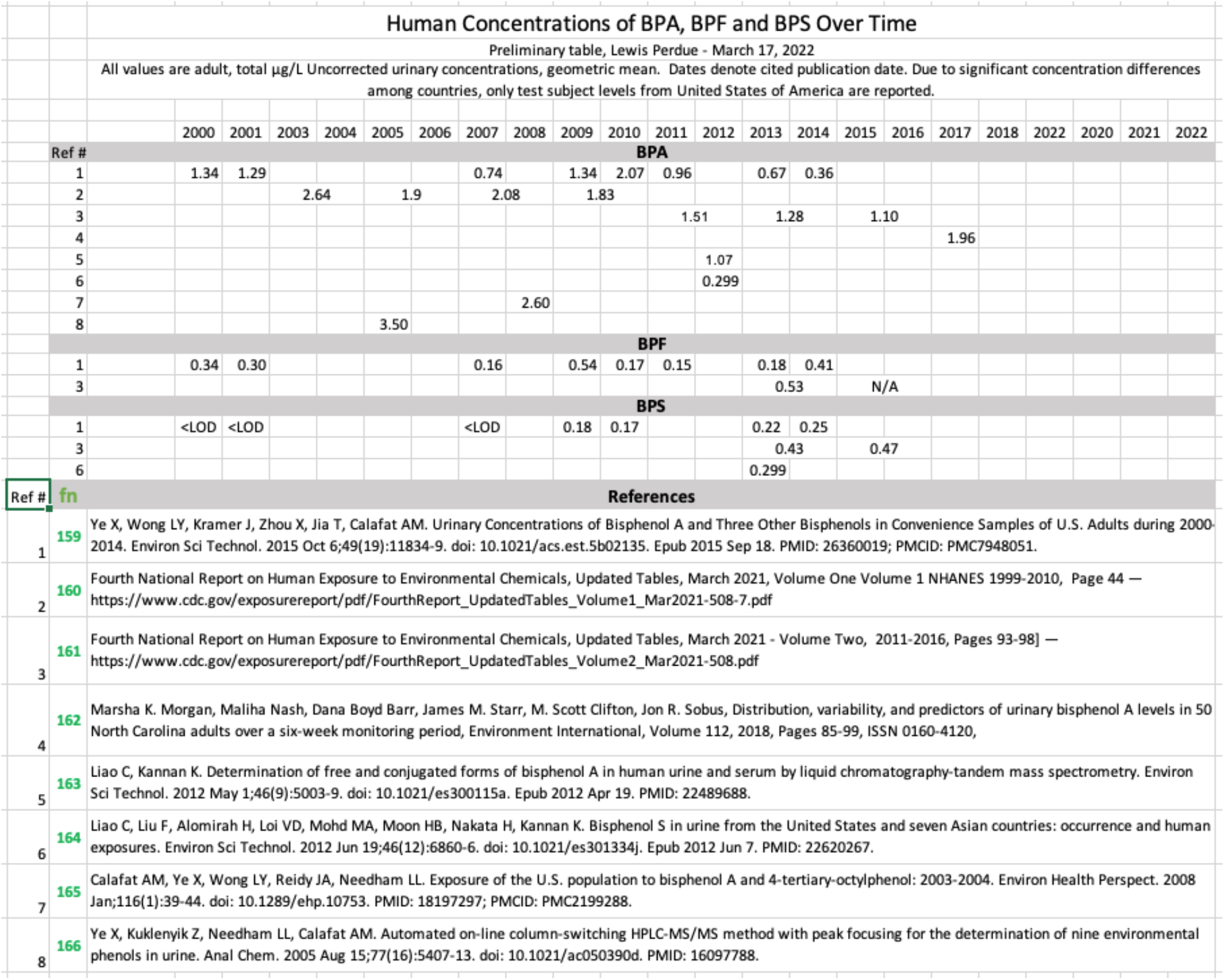

While BPA was not detected, mass spectrometry data indicated that two Bisphenol analogues, Bisphenol F (BPF) and Bisphenol S (BPS) were detected. Those two Bisphenol analogues demonstrated measurable variations that correlated with changes in measured hsCRP, thus lending credence for their sensitivity to Bisphenols.

### Unexpected Result: Missing BPA

In looking at the unexpected absence of BPA, investigators reviewed all the aspects of the study, recipes, ingredients, preparation, and other aspects of the protocols and concluded that the most likely candidate would be a decrease in use of Bisphenol A and the substitution of BPF and BPS in its place: a regrettable substitution. Following that review, investigators believe these results may confirm often-mentioned, but uncited speculation in many published studies that product manufacturers had engaged in regrettable substitutions ^(180-181)^. by reducing BPA usage and substituting Bisphenol analogues which appear to equal or exceed BPA human toxicity^(167-177)^.

However, unwilling to repeat previous speculative statements regarding BPA reduction and substitution, current investigators surveyed peer-reviewed journals seeking indications of that phenomenon. That search yielded no direct data because plastic polymer and additive compositions are closely held corporate secrets, investigators found no direct data for the regrettable substitutions.

As an alternative, investigators sought to determine if historical published data on levels of various Bisphenol levels in human serum could be indicative. Our search criteria focused on human studies in which BPA, BPF and BPS had been measured. Because serum measurements are uncommon and more demanding to conduct, we focused our search on those which had measured BPA levels in urine of residents of the United States.

The vast majority of studies located in the lengthy search were conducted in Asia and the European Union whose citizens consume different foods in often varying ways from people in the United States and whose environmental conditions can vary greatly.

As a result, our starting point of reference was the U.S. Center for Disease Control’s National Health and Nutrition Examination Survey (NHANES) which a program of studies designed to assess the health and nutritional status of adults and children in the United States.

While NHANES studies are conducted infrequently, the Bisphenol data points served as an anchor for additional studies.

Many studies were ruled out because they had not been conducted similar to currently-accepted mass spectrometric procedures, or because results were charted but raw data was not available. As a result, eight published studies measured BPA, but of those, only two measured BPF and three, BPS.

While the sparse results contained in Table 3 are certainly not numerous enough to attain statistical significance, they do appear to indicate that BPA usage is, indeed, decreasing while BPF and BPS are on the increase^(159-166)^. The results are indicative, but not definitive especially when considering the evolution of Bisphenol testing procedures over time and protocols which can vary from lab to lab.

Significantly, we feel that our narrow focus on BPA due to its high visibility, regulatory focus, and its enormous body of published research created a tunnel vision distracted this investigation from a deeper scrutiny of analogues.

Similarly, the practice of regrettable substitutions has also misdirected the focus of other researchers, including CLARITY, and has further obscured scientifically valid efforts to reach regulatory consensus.

#### A Key Lesson In Replicability

Given that dairy products are a major component of the typical American diet, investigators, -- in the early stages of planning explored the possibility of obtaining minimally contaminated raw milk directly from the milking machine, then straining. pasteurizing it, separating the cream, and finally using the resulting milk for direct consumption, butter production and making cheese.

However, by the time appropriate equipment was obtained, investigators found that there were no organic dairies whose herds grazed on pastures not irrigated with recycled municipal wastewater and willing to cooperate. Investigators were forced to conclude that milk/dairy would be non-replicable because of the difficulty of sourcing milk from the same dairy, and that even that effort would fail due to milk variations due to seasonal variations of forage. The same replicability issues also applied to all fresh foods. As mentioned below, the only way to ensure maximum replicability would be to dose the same exact foods used in the contamination and intervention stages with the contaminant being studied.

### Dairy: An Overlooked Clue To Regrettable Substitutions

Regardless of the inherent plastic contamination of milk, the adherence to a typical American diet demanded the inclusion of dairy products. To select dairy products appropriate for the contamination and intervention legs of the study, investigators contracted with Eurofins, a commercial analytic firm with extensive experience in food mass spectrometry to perform LC-MS/MS analyses for the presence of BPA in commercially available milk samples. The Eurofins results produced more unexpected results when data revealed that four of the six samples bought in supermarkets had BPA levels below the LOQ of 0.3 micrograms/Kg. Accordingly, investigators selected the most contaminated sample (1.7 micrograms/Kg) and one below LOQ. Both were selected from national supermarket chains.

In retrospect, the absence of BPA in two-thirds of the samples was a missed clue about BPA’s regrettable substitutions. Testing for BPF in our milk samples was not conducted because, at the time of our investigation, there was little recognition of its potential contamination of dairy^183^. Later published results^184^ confirmed the study’s protocol oversight in overlooking BPF. This oversight was further complicated by investigator’s narrow focus on developing human data to supplement the equivocal data from the CLARITY murine results

### Other Unexpected Results

- Levels of hsCRP unexpectedly decreased during contamination segment. That level was expected to increase. However, intervention level was lower than baseline.
- BPF was approximately the same for Baseline and Intervention.
- BPS was approximately same for all segments.
- BPF-MG was approximately same for baseline and contamination legs. Higher after intervention
- BPS-DG was higher in baseline than contamination. Substantially lower in intervention.

Total serum Bisphenol concentrations offered further confounding results with significant implications for replicability and causality (table 2). Total concentrations of Bisphenols F and S correlated with an increase in the contamination segment and subsequent decrease slightly below baseline for the intervention. This conforms roughly with expectations albeit with an intervention level higher than projected. On the other hand, total monoglucuronide levels showed a decrease during the contamination segment and a return to essentially the same level after intervention.

This may reflect a metabolism phenomenon on which investigators are reluctant to speculate.

As an additional confounding factor. the presence of BPS in serum test results could have resulted from the test subject’s handling of cash register receipts and currency^184^. That issue is addressed, below, in needed measures to control non-dietary environmental exposures

### Limitations

An unfortunate limitation in our experimental design (as well as in Hall et al.^105^) would be the presence of Ultra-Processed Foods (UPFs) in the contamination leg, which may exert similar influences as BPA and its analogues. Complicating the situation further, is the existence of BPA and phthalates in UPFs due to their more extensive plastic contact due to processing and packaging. For these reasons, neither study can conclude whether observed changes are causally due to BPA, UPFs, or both. This situation rendered as irrelevant a further issue of whether nutritional differences in the two legs of the studies could influence the expression of relevant biomarkers. Further, neither this study, nor Hall et al., succeeded in matching the nutritional values of both study legs for calories, total fat, saturated fat, trans fat, sugar, total carbohydrate, dietary fiber, or protein in order to account for possible health effects of those items.

However, even if investigators had been successful, these eight Bisphenol analogues are but a small subset of the 26,000+ biochemicals found so far in foods,^153^ as wells as the 83,000 approved chemicals in the environment -- many of which are metabolically active and could influence clinical biomarkers.

A significant difference between Hall et al. and this trial is that our trial was interrupted by Northern California wildfires that dramatically increased PM2.5 particulate pollution, a notorious promoter of inflammation.^158^ The test subject was exposed to a measurable level of smoke in the process of blood draws That level of exposure may account for the overall smaller decrease in hsCRP in our study versus Hall et al.

## CONCLUSIONS

### Objective 1, validate marker

This trial did reveal a pattern of hsCRP behavior consistent with Hall et al., which measured hsCRP (among numerous other indicators) as an indicator of inflammation in its trial of ultra-processed foods. The sourcing, preparation and serving protocols used in the intervention phase indicated that hsCRP levels appear to have been affected by the trial legs and may serve as a valid indicator.

### Objective 2, evaluate duration

The study protocol’s projections of the metabolic pharmacokinetics of Bisphenol A and hsCRP point to the usefulness of the shortened trial length and indicates that a trial period as short as 6 days could produce useful results

### Objective 3, Verify Causality, and Improve Replication

This trial – like NIH/Hall^105^ and all other published human dietary interventions -- is neither causal nor replicable; conflicts with unknown chemicals and/or additives prohibit a clear relationship with a specific substance as ubiquitous as BPA.

In addition, limiting uncontrollable variables is hugely difficult because basic food substances even from the same source will vary in unknowable ways from month to month, and year to year depending upon weather conditions, water purity, the specific strain of the plant, cultivation, pest control, harvesting, time and conditions for storage, transportation to market, in store, and after purchase.

However, this study developed specific and replicable best practices concerning the training and supervision of kitchen staff along with the sourcing, identification, preparation, and serving of foods, the use of scientific protocols, rigorous data capture, and the control of non-dietary exposures to PDCs that can lead to replicability and causality of subsequent dietary intervention trials involving Plastic-Derived Chemicals.

Our experience showed that for similar studies to be replicable and lead to clinically relevant health recommendations or decisions, such studies must:

a. Apply basic scientific principles and record-keeping,
b. Conduct the study with human subjects,
c. Use exactly the same foods in both legs of the study,
d. Dose foods in the intervention stage using a single compound as an independent variable,
e. Sequester subjects in a disciplined but human-centered dormitory environment to eliminate non-food exposures and other confounding environmental and stress-related psychological confounders.
f. With proper attention to detail, these five requirements could possibly produce clinically useful methods for precision approaches to personalized dietary interventions.^147^

It is worth noting prominently that the use of a dormitory setting by Hall et al is a vital element to control environmental and personal behavior conditions that could bias a dietary intervention study such as those we believe complicated our own investigation. In addition, that study’s extensive set of clinical measurements is invaluable to future investigators seeking human biomarkers for indications of environmental chemical contamination and associated effects.

Initially, investigators did attempt to replicate the dietary portions of Hall et al, but discovered that investigation did not provide menu recipes or nutritional information. In addition, the online menu for the Hall study included many items which involve unknown plastic contact, and possible additives in preparation and packaging^182^.

Full menus, ingredients and nutritional values for this study can be found at the supplemental material online.

This proof-of-concept trial and associated research indicate that dietary intervention studies as a whole are inherently flawed and will not be replicable, causal, or lead to clinically relevant health recommendations or decisions unless they:

While the proof-of-concept study investigation was neither causal nor replicable, the results and knowledge gained merit the conduct of a series of slightly larger (n=10) human trials adhering to the above causality and replicability standards and avoiding the identified confounding factors. If these investigations can be honed to produce causal and replicable results, the protocol could be a time and cost-effective canary in the coal mine protocol that could be required of manufacturers of all chemicals and food additives. The results of these tiny tests would be expanded if sufficient preliminary results indicated reasons for concern.

## Data Availability

Data, including supplemental materials, on request if not already posted and publicly available at
https://stealthsyndromesstudy.com/

## ACKNOWLEDGEMENTS

Investigators are grateful to Ruth N. Muchiri, Ph.D, and Richard B. van Breemen, Ph.D. at the Linus Pauling Institute and College of Pharmacy: Oregon State University for the mass spectroscopy analysis of our human serum samples.

Invaluable assistance was provided by Dr. Roy Gerona, for his lab’s invaluable assistance in providing blood-draw, vacutainer kits, and for initial centrifuging, freezing of samples, and maintaining those at -80C.

We would also like to thank Dr. Alison Abritis Ph.D. for her helpful comments and invaluable discussions and suggestions.

This study was funded by the authors, and the Center for Research on Environmental Chemicals in Humans, Sonoma, CA - https://crechcenter.org/

## SUPPLEMENTAL MATERIAL

Supplemental materials will be posted at https://stealthsyndromesstudy.com/ as they are formatted for upload.

